# Comparison of telephone and in-person interview modalities: duration, richness, and costs in the context of exploring determinants of equitable access to community health services in Meru, Kenya

**DOI:** 10.1101/2024.03.13.24304203

**Authors:** Luke N Allen, Sarah Karanja, John Tlhakanelo, David Macleod, Malebogo Tlhajoane, Andrew Bastawrous

## Abstract

**Background:** Our research team is conducting phenomenological interviews in Kenya with people who have not been able to access community eye health services, aiming to explore the barriers and ideas for potential service modifications. We conducted an embedded study that compared in-person and telephone interview modalities in terms of time requirements, costs, and data richness.

**Methods:** A team of six interviewers conducted 31 in-person interviews and 31 telephone interviews using the same recruitment strategy, topic guide, and analytic matrix for each interview. We compared the mean duration; mean number of themes reported by each participant; total number of themes reported; interviewer rating of perceived richness; interviewer rating of perceived ease of building rapport; number of days taken by the team to complete all interviews; and all costs associated with conducting the interviews in each modality.

**Findings:** In-person interviews were 44% more expensive and took 60% longer to complete than our telephone interviews (requiring 5 days and 3 days respectively). The average in-person interview lasted 110 seconds longer than the average telephone interview (p=0.05) and generated more words and themes. However, the full set of interviews from both approaches identified similar numbers of barriers (p=0.14) and the same number of solutions (p=0.03). Interviewers universally felt that the in-person approach was associated with better rapport and higher quality data (p=0.01). Triangulation of themes revealed good agreement, with 88% of all solutions occurring in both sets, and no areas of thematic dissonance.

**Discussion:** The in-person approach required more time and financial resources, but generated more words and themes per person, and was perceived to afford richer data by interviewers. However, this additional richness did not translate into a greater number of themes that our team can act upon to improve services.

## Introduction

### Background

Qualitative interviews – especially those grounded in the phenomenological approach - are designed to elicit rich data about participants’ lived experiences and perceptions of a given phenomenon.^1^ Our research team has been using interviews to explore barriers to health service access and potential solutions that might improve accessibility by engaging with people who were referred to local eye clinics but were not able to access care in Kenya.^2^

Our project is embedded within the ‘Vision Impact Project’ (VIP) eye screening programme that operates in ten counties across the country. Over a million people have been screened in the past year, and over 150,000 of these people have received care in free local outreach clinics ^3^. However, internal data from the screening programme suggest that up to half of all those referred to local treatment clinics are not accessing care. Furthermore, early data from a related study suggests that certain sociodemographic groups have much lower odds of accessing care than others. In Meru County we have found that younger adults (aged 18-44 years) are the least likely to receive the care they need.^4^ We wanted to explore these peoples’ experiences and perceptions of specific barriers to accessing care, as well as their ideas around any changes we could make to the eye care services to make it easier to access care.

The VIP screening budget is limited, and programme implementers are keen that our interviews can be conducted quickly and as inexpensively as possible whilst still delivering robust findings. The incentive to deliver timely and affordable findings is further underlined by our desire to see embedded qualitative research adopted more widely across routine programmatic quality improvement initiatives, so that the voices of intended service beneficiaries can be included in decision making. Based on the findings of a recent scoping review on rapid qualitative research methods^5^ we have developed a rapid ‘abductive’ interview approach that uses a deductive analytic matrix to facilitate rapid iterative analysis of data whilst “making space for inductive identification of themes and issues not predicted at the outset”.^1^ Our work employs a phenomenological approach, grounded in a pragmatist philosophical paradigm.^6^

The work in Kenya is part of a broader overall project to develop equity-driven and evidence-based approaches to improve access to community-based services across Kenya, Botswana, India and Nepal.^7^ Hundreds of thousands of people are being screened and referred to local services each year, however only around half are able to access care. We wanted to develop an interview approach that could be taken to scale across these four countries – and potentially beyond – to deliver timely insights into how these programmes can be made more accessible, especially for ‘left behind’ groups. Given the scale of the project, telephone interviews are likely to offer the most pragmatic means of obtaining timely insights on how to improve services, however it is not clear what – if anything - would be lost from using telephone interviews as opposed to in-person interviews which generally represent the ‘gold standard’ in qualitative research.^8^

In-person interviews are commonly perceived the ‘best’ way of obtaining rich phenomenological data due to the fact that the interviewer can observe visual cues and quickly build rapport.^9–11^ However, telephone interviews offer unique advantages: the increased social distance can make it easier for participants to discuss sensitive topics; travel time and interviewer safety concerns are eliminated; power imbalances are partially concealed; and overall costs can be greatly reduced – depending on the specific study design and population.^9,12,13^ For our projects, participants can be spread across vast distances, meaning that the risks, costs, and time-requirements for in-person interviews are likely to compare poorly with telephone interviewing. However, we were unable to accurately quantify the trade-off between data quality and resource requirements between the two approaches. As such, we decided to conduct this study to assess which modality offers the best balance of richness, duration, and costs in the context of our work to explore barriers to access and potential solutions in Meru County, Kenya.

### Mode comparison

A number of previous studies have sought to compare telephone and in-person interview modalities.^10,12–17^ In qualitative research, quality is conceptually linked to the ‘richness’ of the data obtained, described by Charmaz in terms of revealing participants’ true feelings, intentions and actions, and accessing their “otherwise inaccessible thoughts”.^18^ Many different proxies have been used to approximate richness in mode comparison studies. A crude but relatively common approach is to measure the duration or wordcount of each interview, working from the assumption that longer interviews, with more words spoken, are more likely to provide deeper insights into people’s lived experiences.^12,13,15,16^ Interview duration is often used in the same way, based on the assumption that longer interviews generate richer data, with some studies also reporting ‘interviewer dominance’ i.e. the proportion of the talking that is done by the interviewer as opposed to the participant.^16^ Surprisingly few qualitative mode effect studies compare the actual content of the interviews, despite the fact that this is a more nuanced way of assessing the amount of topic-related data that are generated.^16,17^ This approach is also relatively straightforward, requiring the reporting of the total number or unique themes that arise from each set of interviews and/or the mean number of themes identified by each interview.

A further approach entails having researchers subjectively rate their experience of each interview in terms of the perceived richness of the data obtained, as done by Abrams et al using a three-point Likert scale.^19^ Other reported measures include quantifying the word count of associated field notes for each interview and counting the amount of conversational turn-taking that occurs in each interview.^15,16^

In this study, we aimed to compare the data richness obtained from two sets of in-person and telephone interviews, electing to use a broad range of proxies: interview duration and wordcount; number of themes identified; and subjective interviewer rating of richness and rapport. We aimed to gather additional data on the time taken to complete each set of interviews, and the associated costs. We hypothesised that telephone interviews would be faster and less expensive to complete than in-person interviews, but offer less-rich data across all metrics of comparison.

## Methods

### Interviews

A previous equity analysis conducted by our team in Meru had found that younger adults (aged 18-44 years old) were the least likely to receive eye care in the county’s community-based screening programme.^4^ We obtained a list of all of the younger adults who did not receive care from Peek Vision, a partner organisation that provides the screening and patient flow management software for the programme.^3^ Peek also record contact numbers for all participants. We used computer-generated random numbers to identify interviewees from this target population, and to determine interview modality.

We performed the telephone interviews first. We performed 36 interviews but a retrospective saturation analysis found that saturation was reached after approximately 30 interviews. Our protocol for this study^20^ stipulated that we would compare an equal number of interviews from both modalities, with a minimum of 20 v 20. We decided to conduct 31 additional in-person interviews and compare these against the first 31 telephone interviews.

The same team of six data collectors conducted all interviews using the same semi-structured interview guide. The same process for audio recording data and directly transcribing quotes into the deductive analytic matrix was used for both modalities, and the same process of iterative review and analysis across all cases within each modality was used to generate the final themes. Data collectors received two days of training before conducting the interviews in September 2023.

### Comparison domains

We gathered data on six domains of richness. We then collated data on the time taken to complete both sets of 31 interviews and their associated costs, following the approach set out in our protocol.^20^

1. **Interview duration**: We measured the duration of each interview in minutes from the start of the consenting process until the researcher concluded the interview e.g. by thanking the participant for answering all of their questions. In line with previous studies discussed above, this metric was used as a proxy for richness, based on the assumption that longer interviews capture richer data than shorter interviews. Note that we did not use interviewer dominance measures this is only possible with typed transcripts, and our approach is based around direct-from-audio entry of verbatim quotes.
2. **Matrix wordcount**: We counted the total number of words entered into the analytic matrix for each set of interviews. These were verbatim quotes directly transcribed from the audio by the data collectors. In line with previous research, we assumed assume that a higher wordcount was associated with richer data.
3. **Total number of themes**: We counted the total number of unique themes for barriers and solutions that were reported across all interviews with each modality. We assumed that the modality that captured the largest number of unique themes was capturing richer data. From an operational standpoint, our underlying study is primarily concerned with generating potential solutions that will improve equitable access, so the number of unique solutions that emerged from each set of interviews is a particularly important metric.
4. **Number of themes reported by each participant**: We also reported the range and mean number of unique themes (barriers and solutions) identified by each participant for each modality. This was to hedge against a situation where one modality generated a greater number of themes than the other, but only because of one or two prolific interviews.
5. **Interviewer subjective rating of richness**: After all of the interviews were complete, each of the six data collectors were asked to provide a single global summary rating of the perceived richness obtained from all in-person and all telephone interviews. Following the approach used by previous researchers, we used a simple Likert scale: low = 1, moderate = 2, high = 3.
6. **Interviewer subjective rating of rapport**: We supplemented the subjective rating of richness with a second question that asked data collectors to provide a global summary rating of the perceived ease of building rapport across all in-person and all telephone interviews. Again, we used a simple Likert scale: low = 1, moderate = 2, high = 3.
7. **Time taken to plan and complete all interviews**: We documented the total amount of time taken to plan and complete all interviews in each modality to the nearest half-day. This was recorded by the Kenyan research manager in charge of scheduling, supervision, and logistics for the local research activities.
8. **Costs**: Working with a health economist, we recorded costs from the payer’s perspective. Both modalities use the same sampling and analytic approach, so we only compared costs that were unique to each approach i.e. those associated with data collection. For telephone interviews these included airtime and staff daily salaries multiplied by the number of days required to complete data collection, starting with the first phone call to recruit the first participant, and ending with the conclusion of the final interview. For in-person interviews we included the costs of printing consent forms, transport for researchers, transport reimbursement offered to participants; payments for local Community Health Promoters and sub-county health officials to assist with setting up the interviews (mobilisation/sensitisation), and staff daily salaries multiplied by the number of days taken for data collection. The costs of voice recorders were not included in the comparison because they were used for both sets of interviews. Similarly, the same two-day training covered skills required for both interview modalities so this was not included in the comparison. We did not compare overhead costs unless they differed for the modalities. The local research manager also recorded any unforeseen additional costs associated with each modality.

### Statistical approach

We used sign tests for the paired interviewer ratings. For the unpaired mean testing comparisons we used histograms to check the data for normality and then used T-tests or Mann-Whitney-U tests, as appropriate, to provide evidence as to whether the two modalities differed across the domains.

### Triangulation of themes

Finally, we compared the barriers and solutions that emerged from both modalities. We identified themes that were identified in both sets of interviews (agreement); themes that emerged from one set of interviews but not the other (silence); and any areas of dissonance i.e. where themes from one set conflicted with those from the other.

### Ethics

Ethical approval was granted by the Kenya Medical Research Institute (KEMRI), the Kenyan National Commission For Science, Technology & Innovation (NACOSTI), and the London School of Hygiene & Tropical Medicine research ethics committee. Each participant provided informed consent.

## Findings

For our comparison we used 31 telephone interviews and 31 in-person interviews that were conducted by our team of six researchers in September 2023 across four sites in Meru County. Table 1 summarises our main findings.

**Table 1:** Performance characteristics of each modality.

| Comparison domain | Metric | Modality |  | p | Ratio |
| --- | --- | --- | --- | --- | --- |
|  |  | In-person | Telephone |  |  |
| **Data richness** | Mean interview duration in minutes and seconds (range) | 10.20 (4.19 - 15.20) | 8.30 (3.10 - 30.10) | 0.005 | 1.11 |
|  | Analytic matrix wordcount – barriers | 4,453 | 2,674 | N/A | 1.67 |
|  | Analytic matrix wordcount – solutions | 2,638 | 2,094 | N/A | 1.26 |
|  | Total number of barriers identified | 15 | 14 | N/A | 1.07 |
|  | Total number of solutions identified | 22 | 22 | N/A | 1.00 |
|  | Mean number of barriers mentioned by each participant (range) | 1.94 (0-4) | 1.58 (0-3) | 0.142 | 1.23 |
|  | Mean number of solutions mentioned by each participant (range) | 2.23 (0-5) | 1.61 (0-4) | 0.029 | 1.39 |
|  | Interviewer global rating of richness (1-3) | 3.0 | 2.0 | 0.014 | 1.5 |
|  | Interviewer global rating of ease of building rapport (1-3) | 3.0 | 2.0 | 0.014 | 1.5 |
| **Time requirement** | Time taken to organise and complete all interviews | 5 days | 3 days | N/A | 1.67 |
| **Costs** | Cost to complete all interviews | USD 668.29 | USD 375.71 | N/A | 1.78 |

**Table 2:** Cost of telephone and physical interviews in USD.

| Line item | In-person | Telephone |
| --- | --- | --- |
| Salaries for two sub-county health officials to schedule in-person interviews | 50.10 | N/A |
| Payments to nine Community Health Promoters for sensitization and mobilization activities | 103.32 | N/A |
| Printing consent forms | 23.29 | N/A |
| Transport for data collectors | 50.10 | N/A |
| Transport reimbursement for interviewees | 97.06 | N/A |
| Airtime | 12.52 | 37.57 |
| Data collector salaries to complete the interviews | 338.15 | 338.15 |
| **Total** | **674.55** | **375.73** |

1 USD = 159.691 KES

### Richness

The average in-person interview lasted 110 seconds longer than the average telephone interview (P=0.05) and generated 33% more words in the analytic matrix. On average, face-to-face interviews identified a greater number of barriers(p=0.14) and solutions (0.03), however the entire in-person interview set only identified one additional barrier and the same number of unique solutions as telephone interviews. All six data collectors were unanimous in their ratings of data richness and ease of building rapport, rating in-person interviews as ‘high’ and telephone interviews as ‘moderate’ for both measures (p=0.01).

### Time requirements

It took two days to prepare for the in-person interviews and then three days to complete them. Preparation time included phoning potential participants to invite them to participate, and then scheduling meeting times and places, and organising transport and local logistics. This included working with local Community Health Promoters (CHPs) and sub-county health officials to sensitise and locate interviewees. This is a vital element in building trust and legitimising our work with participants: the CHPs visited each person to discuss the project and answer any questions, and then supported the researchers to connect them with the interviewees in the field.

#### Costs

Telephone interviews required three days of our data collectors’ time, plus the airtime used to complete the calls. Higher airtime costs for the telephone modality reflects the fact that phone calls were used for recruitment, consenting, and data collection, whereas the in-person approach only used calls for recruitment. Spending on data collectors’ salaries was the same for the in-person interviews – which were also completed in three days - however this modality incurred a number of additional costs. We paid two local county officials to assist with scheduling the in-person interviews, and for sensitization and mobilisation on the days of the interviews. We paid nine Community Health Promoters to build trust, explain the project, and physically locate interviewees. We printed physical consent forms, reimbursed travel to a convenient interview location for our interviewees and paid to transport our data collectors to the same location. Whilst the VIP programme operates across the entire county, at the time that our study was running the programme was operating in Meru town, meaning that all of the interviews were conducted within 15-30 minutes away from our offices. As such, we estimate that the transport costs for data collectors could easily rise by a factor of ten or more for in-person interviews conducted in other parts of the county.

### Triangulation of themes

Table 3 presents the 21 unique barriers that were identified across all 62 interviews. There was agreement between in-person and telephone modalities on nine of these barriers (42.3%). There was silence on the remaining 12 (57.7%) with each modality identifying six unique barriers that did not emerge from the other set of interviews. We found no evidence of thematic dissonance.

**Table 3:** Thematic overlap across in-person and telephone modalities: barriers.

| Barriers | Number of interviews where this barrier was raised |  |
| --- | --- | --- |
|  | In-person | Telephone |
| Conflicting work engagement | 14 | 15 |
| Long queue | 11 | 9 |
| Other conflicting engagement | 7 | 3 |
| Transport costs | 7 | 1 |
| Clinic not open at stated times | 4 | 0 |
| Fear | 3 | 0 |
| Perceived cost of eye drops/spectacles | 2 | 2 |
| Distance to the clinic | 2 | 6 |
| Insufficient numbers of staff | 2 | 2 |
| Lack of clear information on clinic opening times | 2 | 0 |
| Lack of clear information on services available | 2 | 0 |
| Insufficient counselling at the point of referral | 1 | 0 |
| Lack of clear information on the clinic appointment date | 1 | 0 |
| Opportunity costs from loss of wages/income | 1 | 2 |
| Sought services elsewhere | 1 | 2 |
| Forgot | 0 | 3 |
| Did not receive the SMS reminder message | 0 | 2 |
| Assumption that their eye problem wouldn't be addressed | 0 | 1 |
| Dislike of crowded places | 0 | 1 |
| Male health seeking behaviour | 0 | 1 |
| Mixed genders in the queue | 0 | 1 |

Table 4 presents the 25 unique solutions that were identified across all 62 interviews. There was agreement between in-person and telephone modalities on 22 of these barriers (88.0%). There was silence on the remaining three (12.0%) with each modality identifying three unique barriers that did not emerge from the other set of interviews. We found no evidence of thematic dissonance.

**Table 4:** Thematic overlap across in-person and telephone modalities: solutions.

| Solutions | Number of interviews where this barrier was raised |  |
| --- | --- | --- |
|  | In-person | Telephone |
| Hold the clinics on additional days | 10 | 9 |
| Add more staff to each clinic | 9 | 4 |
| Hold the clinics in different locations | 6 | 4 |
| Add a greater number of clinics | 6 | 5 |
| Add more drugs and essential supplies | 4 | 2 |
| Introduce phone call reminders | 4 | 2 |
| Explain the costs and services available | 3 | 0 |
| Extend clinic opening hours | 3 | 2 |
| Specify the clinic dates and times | 3 | 0 |
| Provide a door-to-door service | 2 | 1 |
| Provide transport fare | 2 | 1 |
| Provide transport to the clinic | 2 | 1 |
| Issue public reminders | 2 | 1 |
| Schedule fewer people to attend each clinic every day | 2 | 2 |
| Use SMS reminders | 2 | 2 |
| Have sperate clinic queues for men, women, young & elderly | 2 | 1 |
| Improve staff punctuality | 2 | 1 |
| Hold "mop-up" clinics for those who miss their appointment | 1 | 3 |
| Explain clinic importance at point of referral | 1 | 1 |
| Pay people to attend | 1 | 1 |
| Subsidise treatment | 1 | 0 |
| Hold weekend outreach clinics | 1 | 3 |
| Allow people to choose their appointment day | 0 | 2 |
| Provide specific appointment slots | 0 | 1 |
| Refer non-attenders to the next available clinic via SMS | 0 | 1 |

## Discussion

In this study we examined the quality, costs and time-requirements of in-person vs telephone modes, based on 62 interviews conducted with young adults who had not been able to access eye care services in Meru, Kenya. Even with serendipitously low transport costs, in-person interviews were almost twice as expensive as telephone interviews and took 1.7 times longer to complete. They delivered longer interviews with more words transcribed into the analytic matrix and more themes identified per interview. Our data collectors universally ascribed higher ratings of richness and ease of building rapport to in-person interviews. However, across both modalities, exactly the same number of unique solutions were identified.

Our findings align with the wider literature. Irvine et al. found that telephone interviews tended to be shorter than in-person interviews, although their study only included 11 interviews in total.^15^ Novick’s review of the literature found evidence that telephone interviews are generally less expensive and shorter than in-person interviews.^9^ In their retrospective mode-effect analysis of 300 interviews, Johnson et al. found that in-person interviews produced longer transcripts and more word-dense field notes, but generated the same themes as telephone and videocall-based interviews.^16^ Interestingly, subjective interviewer ratings were also similar across the approaches. Krouwel and colleagues also compared in-person interviews to those conducted using video-calling software. They found that in-person interviews generated more data but the overall number of themes derived from each approach was similar.^17^ Vogl’s triangulation of the themes that emerged from two sets of interviews with 56 children found negligible differences.^13^ Finally, in his systematic review comparing telephone and in-person approaches, Rahman concludes that both telephone and in-person modalities can generate comparably rich data, with telephone interviews tending to be less time consuming and less expensive.^10^

Given that empirical mode comparisons consistently find that remote interviews are able to generate similar qualitive themes at lower costs and in shorter time periods than in-person interviews, irrespective of research question and population studied, Rahman has argued that the in-person modality should only be used if the specific research question demands it.^10^ The relationship between depth of detail, number of themes, and agreement between themes is intriguing. Participants tend to provide much more detail about a given phenomena during in-person interviews, as indicated by longer transcripts, interview durations, and analytic matrix wordcounts. However, this additional detail rarely translates into identification of novel codes or themes when compared to remote approaches.

There were differences in the themes that emerged from both sets of interviews. Whilst the differences between the solutions was fairly minor, several of the barriers that were raised during the telephone interviews were potentially more candid than those derived from in-person interviews. A form of social desirability bias might have been at play, with interviewees feeling more comfortable disclosing potentially embarrassing or taboo issues when the interviewer was not physically sat in front of them.^21,22^ Some of the barriers that emerged exclusively from the phone interviews included forgetting about the appointment, assuming that the service would not meet their needs, and perceiving the mixing of men and women in a single queue as ‘shameful’.

In terms of strengths and limitations, whereas most research in this field tends to employ one or two metrics, our study compared eight different dimensions of performance, including proxies for richness (duration and wordcount), mean and aggregate themes, and subjective interviewer ratings, supplemented with an assessment of costs and time requirements. We conducted a relatively large number of interviews on a topic that is central to global efforts to extend Universal Health Coverage as part of the Sustainable Development Goals.^23–25^

Our sample size was based on a post-hoc saturation analysis, but the decision to compare 31 interviews with each modality rather than 30 vs 30 was essentially arbitrary. The generalisability of our findings is limited by our relatively focused research question and the homogeneity of our population (younger adults in Meru who were found to have an eye problem during screening but did not manage to access further care). Ultimately, whilst our study presents multiple measures we are not able to definitely say which approach is best, as there is no single ‘right’ way to balance differences in richness, costs, and time requirements.

Previous research has documented that the impact of qualitative research findings on real-world programmes is influenced by the timeliness of the findings,^26,27^ and our broader embedded qualitative work places a premium on rapidly identifying barriers and potential solutions to improve equitable access to care within a live, ongoing screening programme. Given our focus on identifying solutions and service modifications that can be rapidly tested, the lack of dissonance between the modes, lower costs, lower time requirements, and additional researcher safety benefits associated with telephone interviews means that we are very likely to continue using this approach.

## Conclusions

Our set of 31 telephone interviews was completed in less time and at less expense than the 31 in-person interviews. Whilst the in-person modality generated longer interviews and more data, the ultimate number of themes that derived from both sets was nearly identical. For our purposes, telephone interviews offer clear operational advantages with no meaningful reduction in data quality.

## Data Availability

Data produced in the present study are available upon reasonable request to the authors

## Declarations

### Author contributions

LA conceived the study with SK. LA and SK performed the data collection and analyses. LA wrote the initial draft. SK and MT provided additional written content. JT provided health economics input and reviewed the manuscript. DM provided statistical input and reviewed the manuscript. AB reviewed the protocol and provided additional methodological comments. All authors reviewed and approved the final version of the manuscript.

### Ethical approval

The study was granted ethical approval by KEMRI scientific and ethics review unit on November 30^th^ 2022, NACOSTI on 12^th^ January 2023, and the LSHTM ethics committee on 5^th^ May 2023.

## References

1. Pope C, Mays N. Qualitative Research in Health Care, 4th Edition | Wiley [Internet]. Oxford: Wiley Blackwell; 2020 [cited 2023 Jan 5]. Available from: https://www.wiley.com/en-gb/Qualitative+Research+in+Health+Care%2C+4th+Edition-p-9781119410836

2. Allen LN, et al. Improvement Studies for Equitable and Evidence-based Innovation: an overview of the ‘IM-SEEN’ model. Int J Equity Health. 2023;In Press.

3. Peek Vision. News [Internet]. Kenya’s Vision Impact Project reaches one million people in its first year. 2018 [cited 2023 Oct 13]. Available from: https://peekvision.org/en_GB/news/

4. Allen L, Karanja S, Gichangi M, Bunywera C, Rono H, Macleod D, et al. Access to community-based eye services in Meru, Kenya: a cross-sectional equity analysis [Internet]. medRxiv; 2024 [cited 2024 Mar 1]. p. 2024.02.23.24303185. Available from: https://www.medrxiv.org/content/10.1101/2024.02.23.24303185v1

5. Allen LN, Azab H, Jonga R, Gordon I, Karanja S, Thaker N, et al. Rapid methods for identifying barriers and solutions to improve access to community health services: a scoping review. BJGP Open. 2023 Jul 19;0047.

6. Legg C, Hookway C. Pragmatism. In: Zalta EN, editor. The Stanford Encyclopedia of Philosophy [Internet]. Summer 2021. Metaphysics Research Lab, Stanford University; 2021 [cited 2022 May 3]. Available from: https://plato.stanford.edu/archives/sum2021/entries/pragmatism/

7. Allen LN, Nkomazana O, Mishra SK, Gichangi M, Macleod D, Ramke J, et al. Improvement studies for equitable and evidence-based innovation: an overview of the ‘IM-SEEN’ model. Int J Equity Health. 2023 Jun 17;22(1):116.

8. Azad A, Sernbo E, Svärd V, Holmlund L, Björk Brämberg E. Conducting In-Depth Interviews via Mobile Phone with Persons with Common Mental Disorders and Multimorbidity: The Challenges and Advantages as Experienced by Participants and Researchers. Int J Environ Res Public Health. 2021 Nov 11;18(22):11828.

9. Novick G. Is There a Bias Against Telephone Interviews In Qualitative Research? Res Nurs Health. 2008 Aug;31(4):391–8.

10. Rahman RBA. Comparison of Telephone and In-Person Interviews for Data Collection in Qualitative Human Research. Interdiscip Undergrad Res J [Internet]. 2023 Mar 5 [cited 2023 Dec 7]; Available from: https://indigo.uic.edu/articles/journal_contribution/Comparison_of_Telephone_and_In-Person_Interviews_for_Data_Collection_in_Qualitative_Human_Research/22217215/1

11. Rubin HJ, Rubin IS. Qualitative Interviewing: The Art of Hearing Data. SAGE; 2011. 289 p.

12. Sturges JE, Hanrahan KJ. Comparing Telephone and Face-to-Face Qualitative Interviewing: a Research Note. Qual Res. 2004 Apr 1;4(1):107–18.

13. Vogl S. Telephone Versus Face-Toface Interviews: Mode Effect on Semistructured Interviews with Children. Sociol Methodol. 2013;43:133–77.

14. Francis JJ, Johnston M, Robertson C, Glidewell L, Entwistle V, Eccles MP, et al. What is an adequate sample size? Operationalising data saturation for theory-based interview studies. Psychol Health. 2010 Dec;25(10):1229–45.

15. Irvine A, Drew P, Sainsbury R. ‘Am I not answering your questions properly?’ Clarification, adequacy and responsiveness in semi-structured telephone and face-to-face interviews. Qual Res. 2013 Feb 1;13(1):87–106.

16. Johnson DR, Scheitle CP, Ecklund EH. Beyond the In-Person Interview? How Interview Quality Varies Across In-person, Telephone, and Skype Interviews. Soc Sci Comput Rev. 2021 Dec 1;39(6):1142–58.

17. Krouwel M, Jolly K, Greenfield S. Comparing Skype (video calling) and in-person qualitative interview modes in a study of people with irritable bowel syndrome – an exploratory comparative analysis. BMC Med Res Methodol. 2019 Nov 29;19(1):219.

18. Charmaz K, Henwood K. Grounded Theory. In Smith J. A. (Ed.), Qualitative psychology: A practical guide to research methods [Internet]. SAGE Publications Ltd; 2003 [cited 2024 Jan 11]. 81–110 p. Available from: https://methods.sagepub.com/book/the-sage-handbook-of-qualitative-research-in-psychology

19. Abrams KM, Wang Z, Song YJ, Galindo-Gonzalez S. Data Richness Trade-Offs Between Face-to-Face, Online Audiovisual, and Online Text-Only Focus Groups. Soc Sci Comput Rev. 2015 Feb 1;33(1):80–96.

20. Allen L, Karanja S, Tlhajoane M, Tlhakanelo J, Macleod D, Bastawrous A. A protocol for the comparison of telephone and in-person interview modalities: duration, richness, and costs in the context of exploring determinants of equitable access to community health services in Meru, Kenya. medRxiv [Internet]. 2024; Available from: https://www.medrxiv.org/content/10.1101/2024.03.04.24303701v1

21. Bispo Júnior JP. Social desirability bias in qualitative health research. Rev Saude Publica. 2022;56:101.

22. Kreuter F, Presser S, Tourangeau R. Social Desirability Bias in CATI, IVR, and Web Surveys: The Effects of Mode and Question Sensitivity. Public Opin Q. 2008 Dec 1;72(5):847–65.

23. World Health Organization. Thirteenth general programme of work, 2019–2023: promote health, keep the world safe, serve the vulnerable [Internet]. World Health Organization; 2019 [cited 2021 Nov 11]. Report No.: WHO/PRP/18.1. Available from: https://apps.who.int/iris/handle/10665/324775

24. World Health Organization. Universal health coverage (UHC) [Internet]. 2021 [cited 2021 Nov 11]. Available from: https://www.who.int/news-room/fact-sheets/detail/universal-health-coverage-(uhc)

25. UN General Assembly. A/RES/70/1: Transforming our world: the 2030 Agenda for Sustainable Development [Internet]. 2015 Sep [cited 2021 Nov 11]. Available from: https://www.un.org/ga/search/view_doc.asp?symbol=A/RES/70/1&Lang=E

26. Johnson GA, Vindrola-Padros C. Rapid qualitative research methods during complex health emergencies: A systematic review of the literature. Soc Sci Med. 2017 Sep 1;189:63–75.

27. Allen LN, Azab H, Jonga R, Gordon I, Karanja S, Thaker N, et al. Rapid methods for identifying barriers and solutions to improve access to community health services: a scoping review. BJGP Open [Internet]. 2023 Dec 1 [cited 2024 Feb 8];7(4). Available from: https://bjgpopen.org/content/7/4/BJGPO.2023.0047

